# Association of Animal Ownership and Short-Term Rainfall Events with under-5 Diarrhea Prevalence in sub-Saharan Africa

**DOI:** 10.64898/2025.12.22.25342865

**Authors:** Steven Sola

## Abstract

According to the 2021 Global Burden of Diseases Study, diarrhea contributed to an estimated 340,429 deaths among children younger than five years old and an estimated 245,966 deaths in sub-Saharan Africa specifically. Some efforts to reduce the burden of diarrhea in Africa have focused on animal fecal contamination, particularly during extreme precipitation events (EPEs). A focus has been the concentration-dilution hypothesis, which is centered on EPEs immediately following dry periods.

The source population included 12,007,303 individuals from 172 surveys between 1990 and 2022. We conducted descriptive analyses and modeled associations between EPEs and diarrheal prevalence on a study population that included 413,576 children under five whose families owned livestock. EPEs that occurred 15-21 days and 22-28 days before the surveys were administered showed a statistically significant increase in diarrhea prevalence compared to EPEs not occurring during those time periods. We utilized a mixed-effects Poisson regression model with a log link to assess the association between individual animal species and the two-week prevalence of diarrhea among children under the age of 5. There was a decreased two-week prevalence of diarrhea among families that own horses or donkeys and were exposed to an EPE (PR: 0.87, 95% CI: 0.83-0.92, p-value: <0.001), but an increased two-week prevalence among families that own pigs and experienced an EPE (PR: 1.15, 95% CI: 1.03-1.29, p-value: 0.01) compared to families that did not own these animals and did not experience an EPE.

The results from this study further illustrate the complex relationship between the occurrence of EPEs and the prevalence of diarrhea. Future studies can refine the use of EPEs in their analysis to look at the occurrence of EPEs immediately after local droughts. Animal husbandry practices are also vital to improve the understanding of their role in diarrheal outcomes for the family.

## Introduction

According to the 2021 Global Burden of Diseases Study, diarrhea contributed to an estimated 340,429 deaths among children younger than five years old and an estimated 245,966 deaths in sub-Saharan Africa [1]. Despite a trend of decreasing deaths due to diarrheal diseases, sub-Saharan Africa has among the highest rates of diarrheal disease [1]. Some of these diarrheal deaths are the result of zoonotic infections, typically related to animal exposure. While animals are a source of potentially deadly diarrheal pathogens, they also can be a source of income and nutrition to families, especially those from a lower socioeconomic class [2,3]. Given the need to reduce the burden of diarrheal diseases among these populations, especially in sub-Saharan Africa, more clear understanding of the contributions of peridomestic animals is needed, especially in an ever-evolving climate change scenario.

A “concentration-dilution hypothesis” has been proposed by Levy et al [4]. and Carlton et al [5]. which is focused on the build-up of pathogens in the environment during dry conditions, which are subsequently released into the environment during heavy rainfall events (thus delivering a concentrated amount) (see figure 1). However, during wet periods, these pathogens don’t have time to concentrate in the environment, thus diluting the potential exposure during any one period. A systematic review and metanalysis by Kraay et al [6]. shows support for this hypothesis, suggesting that rainfall patterns, alone or in combination with temperature, may impact the relationship between peridomestic animal keeping and health outcomes, particularly in vulnerable populations like children under five years old.

**Figure 1.**
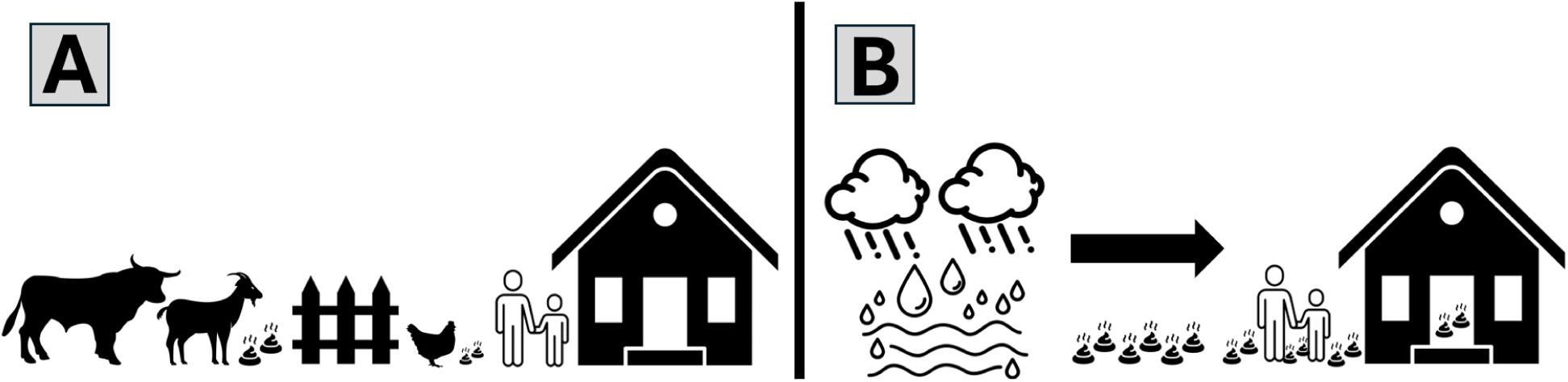
An illustration of the concentration-dilution hypothesis. In Part A, bigger animals, such as bulls and goats, are usually housed in areas away from the home, typically behind a fence. Meanwhile, smaller animals, such as chickens, are typically left to roam freely near the house and compound. During dry periods, feces from these animals may build up in the environment, unless they are actively disposed of by the owners. In Part B, whenever there is a precipitation event, and especially an extreme precipitation event (EPE), these feces are dispersed throughout the immediate environment, leading to an increase in human-animal fecal exposures.

Our prior work on animal ownership showed that households that owned animals had a higher diarrhea prevalence, but at the same time, households that owned more than one animal species had a lower diarrhea prevalence compared to households that only owned one species of animal. However, our prior work also identified that socioeconomic status, which can be correlated with animal ownership, was important. Due to the several climate zones throughout this subcontinent, we planned to research the interaction between animal ownership and short-term rainfall events (STRE) on the prevalence of diarrhea among children under the age of five in sub-Saharan Africa. This analysis combined survey and weather data together to further our understanding of the role of local weather patterns on diarrheal outcomes and the potential mechanisms behind the concentration-dilution hypothesis. The hypothesis for this research is that, among households with peridomestic animal keeping, children exposed to an STRE will have a greater two-week prevalence of diarrhea compared to children that are not exposed to an STRE.

## Methods

### Study Design

To estimate the association between animal ownership, extreme precipitation events, and under-5 diarrhea, we leveraged the United States Agency for International Development’s (USAID) Demographic and Health Surveys (DHS) data, similar to prior work. These data were combined with ERA5-Land weather data from the European Centre for Medium-Range Weather Forecasts (ECMWF) [7] in order to generate lagged interval and cumulative rainfall patterns for households included in the DHS surveys from 2005 to 2022.

### Survey Data

For this analysis, our exclusion criteria were: all individuals who lived in urban or unknown urbanicity settings, according to DHS definitions [8]. We also excluded everyone five years of age and older, those who had a missing age, or children who were already deceased.

Additionally, we excluded children whose families did not own any animals or those who did not have any animal ownership information. Finally, we excluded households that had unrealistic GPS coordinates, such as households located at a latitude and longitude of (0,0). We used this final dataset to conduct our descriptive and mixed-effects model results.

### Weather Data

ERA5-Land is a climate reanalysis dataset at the spatial resolution of 0.1’ x 0.1’ (approximately 9 kms x 9 kms). A total of 6,573 files of daily data (January 1, 2005 – December 31, 2022) were downloaded in NetCDF format from ECMWF. The primary variable of interest for this study was total precipitation (tp). Köppen-Geiger Zones (KGZ) were recorded for each household based on their GPS coordinates to the nearest 100 decimal seconds using the *kgc* R package [9].

### Statistical Analysis

We utilized a mixed-effects Poisson regression model with a log link to assess the association between individual animal species and the two-week prevalence of diarrhea among children under the age of 5. The results of these models are prevalence ratios. We utilized the *lme4* package in R [10]. Our model was specified as follows:

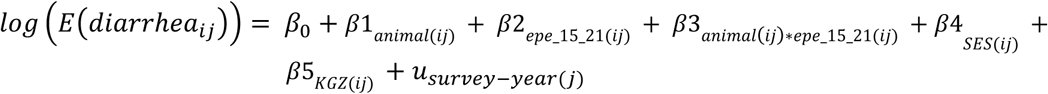

Where:

β_1_ is to the specific animal of interest (Poultry [Chicken/Duck]), Bull/Cow/Cattle, Goat/Sheep, Horse/Donkey, Pig).

β_2_ is the presence or absence of an STRE in the 15-21 days preceding the administration of the survey (dichotomous).

β_3_ is the interaction term between each individual animal and the presence of an short-term rainfall event (STRE).

β_4_ is to the SES of the household that the individual lives in (ordinal).

β_5_ is to the Köppen-Geiger Zone (KGZ) of the household that the individual lives in (categorical).

*u* is the random intercept for the survey-year (e.g. “South Africa 2016” or “Senegal 2007”).

subscript *i* is used to denote measurements that change on the individual level.

subscript *j* is used to denote measurements that change on the group (i.e. “survey-year”) level.

The primary outcome for our analysis was the variable “H11”, which questions whether the child has had diarrhea within the past two weeks or 24 hours. We combined these questions so that all children with diarrhea were recorded as having a two-week diarrhea prevalence. The primary exposures for this analysis are the species of animals that a family owns, and whether an STRE occurred at the family’s location in the 15-21 days before the survey administration. We were interested in 15-21 days before the survey administration because there was a 2-week recall in the survey, and most diarrheal pathogens have a short incubation timeframe. We created an interaction term between each species of animal that the family owns and the presence, or absence, of an STRE in the 15-21 days before the survey administration. Using this interaction term, we assessed the multiplicative joint effect of this interaction term by adding the fixed effects estimates for β_1_, β_2,_ and β_3_ and exponentiating the result. The result of this was the two-week diarrhea prevalence of a child whose family owned each species and experienced an STRE in the 15-21 days before the administration of the survey, compared to the two-week diarrhea prevalence of a child whose family did not own each species and did not experience an STRE in the 15-21 days before the administration of the survey.

Information about animal ownership (HV246) and SES (HV270) were extracted from each survey included in the analysis. As shown in our previous work using the same source population as this one, SES was included in this model because two-week prevalence of diarrhea was shown to be differential across the different SES levels. KGZ was included in our model as rainfall levels and extreme precipitation events are dependent on the particular climate characteristics of each area. Including KGZ into the model can also help us account for seasonality for each household. We accounted for clustering at the survey-year level by entering that term as a random intercept into the model. We considered the inclusion of random intercepts for household and cluster, but ultimately excluded these as they did not significantly change the results from the model, and to keep the model parsimonious.

All analyses were completed using R 4.2.1 [11]. Replication code is made publicly available on https://github.com/sqsola/ClimateWASH. Data can be obtained through a request to the Demographic and Health Survey team at https://dhsprogram.com, although this is subject to change. Permission to download data was received prior to commencing data analyses.

### Short-Term Rainfall Events and Time Lags

We assessed the association between STREs and the two-week prevalence of diarrhea among children under the age of five in a household. We defined STRE as a single day of precipitation that was above the 95^th^ percentile of the total precipitation of the time period. If a household was located in an area where the seven-day total precipitation was two millimeters, then the 95^th^ percentile would be 1.9 millimeters, and a household would be exposed to an STRE if a single day within that seven-day period had a precipitation of 1.9 millimeters. We used a percentile method as the definition of STREs based on the definitions used in previous research [6,12–15]. Additionally, we conducted sensitivity analyses using the 99^th^ and 99.9^th^ percentiles as the definition of an STRE within our mixed-effects Poisson regression models.

To assess the association between STREs and prevalence of diarrhea, we investigated when those STREs occurred before the date of the survey. We investigated the following time lags in this study: 15-21 days, 15-28 days, 22-28 days, 30-60 days, and 15-60 days (see figure 2). Given the uncertainty of when a child’s diarrheal outcome started before administration of the survey, we also considered the incubation period for common diarrheal pathogens, such as rotavirus, *G. lamblia*, and *E. coli* (see supplemental table 2, based on a study by Thystrup et al. [16]). We did not assess any time lags within the two weeks of the potential recall period for diarrhea to avoid issues related to diarrhea occurring before an STRE (i.e. to avoid introduction of reverse causation in which the STRE occurred after the reported diarrheal event during the two-week recall period).

**Figure 2.**
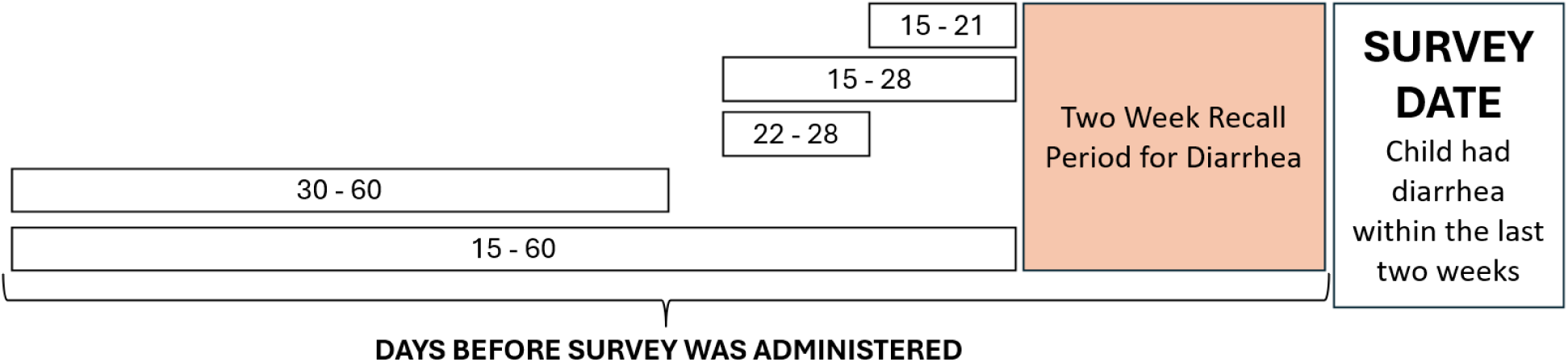
Time lags used in this study. Lags are the period of time before the administration of the survey. These surveys use a two-week diarrhea prevalence, so time lags were only considered for the time before the two-week diarrhea recall period.

## Results

Our source dataset included 12,007,303 individuals across 172 survey-years. Exclusion criteria were: all individuals who lived in urban (n = 3,116,550) or unknown urbanicity (n = 1,747,960) settings, according to DHS definitions [8]. We also excluded everyone five years of age and older (n = 1,614,644), those who had a missing age (n = 4,742,118), or children who were already deceased (n = 259). We further excluded children whose families did not own any animals (n = 173,530), or those who did not have any animal ownership information (n = 193,347). Finally, we excluded households that had unrealistic GPS coordinates, such as those with a GPS of (0,0) (n = 5,413). A total of 413,576 children aged four years or younger whose household owned at least one animal (see figure 3). Our dataset encompassed 99 survey-years from 35 countries (see supplemental table 1).

**Figure 3.**
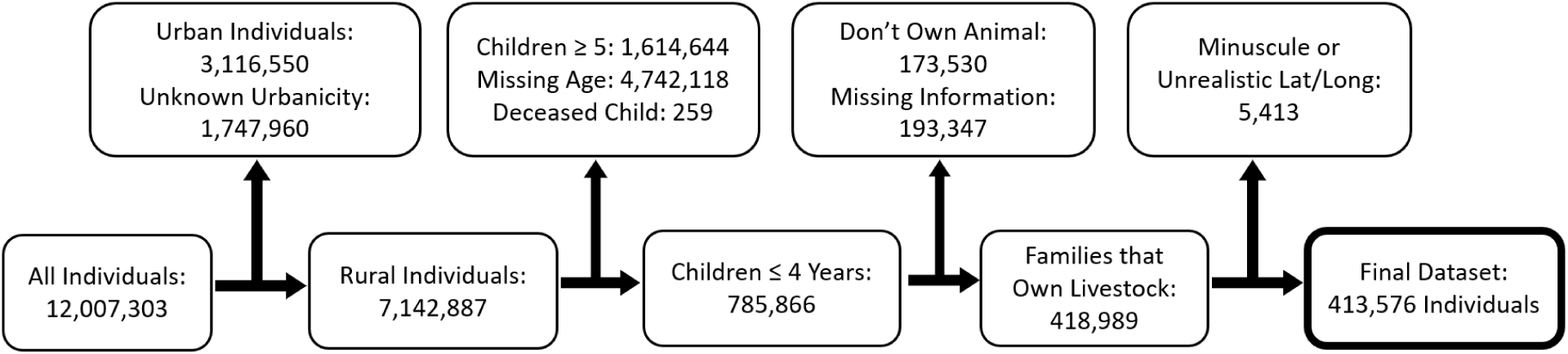
Inclusion of Individuals in Analysis

### Descriptive Analyses

The locations of each household included in this analysis and their associated KGZ can be seen in Figure 4.

**Figure 4.**
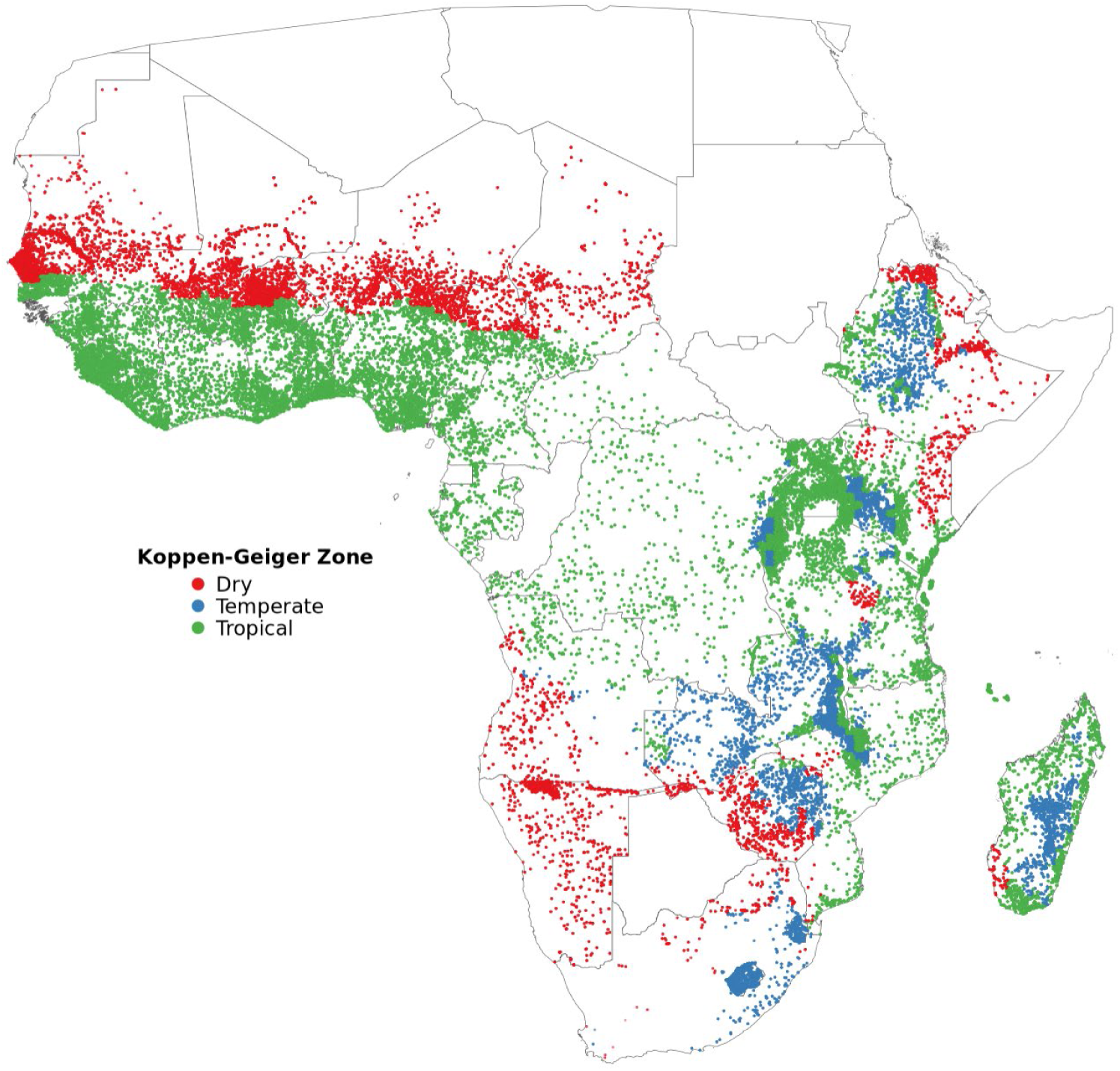
Households included in the study and their associated Koppen-Geiger Climate Classification Zone

There were 128,148 children located in a dry KGZ, 58,303 children located in a temperate KGZ, and 227,125 children located in a tropical KGZ. The average 7-day rainfall for the dry, temperate, and tropical KGZ were 0.83 cm, 2.93 cm, and 3.08 cm, respectively. There was also a difference in the distribution of animals across the KGZs. Poultry (chicken/ducks) were the most owned category of animal (76.1% of total households), followed by goats/sheep (56.4% of total households). Households in the dry KGZ had the highest percentage ownership of goat/sheep, bulls/cows/cattle, and horses/donkeys, while households in tropical KGZ had the highest household ownership of chickens (80.8%). Households in Dry KGZ had a much higher ownership of horses and donkeys compared to households in temperate and tropical KGZs (53.7% compared to 11.5% and 8.7%, respectively), as well as goats and sheep (73.0% compared to 36.4% and 35.7%, respectively). Alternatively, dry KGZ households had less pig ownership compared to these two other KGZs (see table 1).

**Table 1.** Köppen-Geiger Zones and the number of children that live within each zone. Also included is the average 7-day rainfall in each zone, and the number of children whose households own the five categories of animals of interest in this research.

| Köppen-Geiger Zone | Number of Children | Average 7-Day Rainfall (cm) | Poultry (chicken/ducks) | Goat/Sheep | Bull/Cow/Cattle | Horse/Donkey | Pig |
| --- | --- | --- | --- | --- | --- | --- | --- |
|  |  |  | (n, %) | (n, %) | (n, %) | (n, %) | (n, %) |
| Dry | 128,148 | 0.83 | 88,421 (69.0%) | 93,529 (73.0%) | 72,879 (56.9%) | 68,859 (53.7%) | 3,437 (2.7%) |
| Temperate | 58,303 | 2.93 | 42,756 (73.3%) | 21,230 (36.4%) | 28,175 (48.3%) | 6,705 (11.5%) | 8,709 (14.9%) |
| Tropical | 227,125 | 3.08 | 183,471 (80.8%) | 118,592 (52.2%) | 81,060 (35.7%) | 19,841 (8.7%) | 24,618 (10.8%) |
| Total | 413,576 |  | 314,648 (76.1%) | 233,351 (56.4%) | 182,114 (44.0%) | 95,405 (23.1%) | 36,764 (8.9%) |

There were five time lags that had statistically significant differences between whether a child experienced an STRE: 15-21 days and 22-28 days. Of these, the 22-28 day time lag had the greatest difference between those children that did and did not experience an STRE. The time lags of 15-28 days and 30-60 days were characterized by children having a higher diarrhea prevalence when they weren’t exposed to an STRE compared to children that were exposed to an STRE. Children that were and weren’t exposed to an STRE had an almost equal diarrhea prevalence in the 15-60 day time lag (see table 2). All the time lags showed a statistically significant difference in the SES levels between those that did and did not experience STREs, with those that did not experience STREs having a higher SES compared with those that did not experience STREs. There was no statistically significant difference in the average ages of children that did and did not experience STREs.

**Table 2.** STREs and diarrhea prevalence, along with their associated SES levels and ages.

| Short-Term Rainfall Event (STRE)? | Children Under 5 with Animals | Children Under 5 with Animals and Diarrhea | Diarrhea Prevalence | p-value<sup>1</sup> | Average SES<sup>2</sup> | p-value<sup>3</sup> | Average Age | p-value<sup>3</sup> |
| --- | --- | --- | --- | --- | --- | --- | --- | --- |
| 15-21 Days Before Survey (one week before recall period) |  |  |  |  |  |  |  |  |
| No | 373,047 | 45,439 | 12.18 | < 0.001 | 2.27 | < 0.001 | 1.97 | 0.08 |
| Yes | 40,529 | 5,298 | 13.07 |  | 2.07 |  | 1.99 |  |
| 15-28 Days Before Survey (two weeks before recall period) |  |  |  |  |  |  |  |  |
| No | 394,667 | 48,440 | 12.27 | 0.61 | 2.26 | < 0.001 | 1.97 | 0.59 |
| Yes | 18,909 | 2,297 | 12.15 |  | 1.95 |  | 1.98 |  |
| 15-60 Days Before Survey |  |  |  |  |  |  |  |  |
| No | 412,242 | 50,573 | 12.27 | 0.98 | 2.25 | < 0.001 | 1.97 | 0.55 |
| Yes | 1,334 | 164 | 12.29 |  | 1.94 |  | 2.00 |  |
| 22-28 Days Before Survey |  |  |  |  |  |  |  |  |
| No | 371,733 | 45,031 | 12.11 | < 0.001 | 2.27 | < 0.001 | 1.97 | 0.16 |
| Yes | 41,843 | 5,706 | 13.64 |  | 2.09 |  | 1.98 |  |
| 30-60 Days Before Survey |  |  |  |  |  |  |  |  |
| No | 411,394 | 50,474 | 12.27 | 0.76 | 2.25 | < 0.001 | 1.97 | 0.74 |
| Yes | 2,182 | 263 | 12.05 |  | 1.94 |  | 1.98 |  |

<sup>1</sup>Two-proportions Z-test
<sup>2</sup>SES is an ordinal variable with the following levels: 1 = Poorest, 2 = Poor, 3 = Middle Class, 4 = Rich, 5 = Richest
<sup>3</sup>Mann-Whitney U-test

### Modeling Analyses

After conducting our mixed-effects Poisson regression with a log link, we found that there were two models that showed a statistically significant difference in the prevalence of two-week diarrhea when examining the animals that a household owned (see table 3). One model was for households that owned bulls, cows, or cattle. Children in these households had an increased prevalence of two-week diarrhea compared to children whose families did not own bulls, cows, or cattle (PR: 1.02, 95% CI: 1.00-1.04, p-value 0.04). The other model was for households that owned horses or donkeys, and children in these households had a decreased prevalence of two-week diarrhea compared to households that did not own horses or donkeys (PR: 0.95, 95% CI: 0.92-0.98, p-value: <0.001). The bull/cow/cattle model showed a statistically significant increase in the prevalence of two-week diarrhea among children in tropical climate zones compared to children in dry climate zones. The model where horses were the primary exposure showed a statistically significant decrease in the prevalence of two-week diarrhea among children living in temperate climate zones compared to children living in dry climate zones (PR: 0.96, 95% CI: 0.92-1.00, p-value: 0.03). All five models showed a statistically significant decrease in the prevalence of two-week diarrhea among children that were a part of richer SES groups compared to children that were a part of the poorest SES group.

**Table 3.** Model results from the mixed-effects Poisson regression models with a log link. The outcome of these models is a prevalence ratio of children with two-week diarrhea compared to children that did not experience two-week diarrhea. An STRE was a single day of precipitation that was above the 95^th^ percentile of the total precipitation of the time period. The joint effect from the interaction term is the prevalence of the 2-week reported diarrhea when considering each animal along with the presence of an STRE compared to the prevalence of the 2-week reported diarrhea when considering the absence of each animal in the absence of an STRE.

|  | Poultry (Chickens/Ducks) |  |  | Bull/Cow/Cattle |  |  | Goat/Sheep |  |  | Horse/Donkey |  |  | Pig |  |  |
| --- | --- | --- | --- | --- | --- | --- | --- | --- | --- | --- | --- | --- | --- | --- | --- |
|  | PR | 95% CI | p-value | PR | 95% CI | p-value | PR | 95% CI | p-value | PR | 95% CI | p-value | PR | 95% CI | p-value |
|  | 1.00 | 0.98, 1.02 | >0.9 |  |  |  |  |  |  |  |  |  |  |  |  |
|  |  |  |  | 1.02 | 1.00, 1.04 | **0.04** |  |  |  |  |  |  |  |  |  |
|  |  |  |  |  |  |  | 1.00 | 0.98, 1.02 | >0.9 |  |  |  |  |  |  |
|  |  |  |  |  |  |  |  |  |  | 0.95 | 0.92, 0.98 | **<0.001** |  |  |  |
|  |  |  |  |  |  |  |  |  |  |  |  |  | 0.99 | 0.96, 1.02 | 0.5 |
| **Köppen-Geiger Zone** |  |  |  |  |  |  |  |  |  |  |  |  |  |  |  |
| Dry | Ref | Ref |  | Ref | Ref |  | Ref | Ref |  | Ref | Ref |  | Ref | Ref |  |
| Temperate | 0.97 | 0.93, 1.01 | 0.12 | 0.97 | 0.93, 1.01 | 0.10 | 0.97 | 0.93, 1.01 | 0.12 | 0.96 | 0.92, 1.00 | **0.027** | 0.97 | 0.93, 1.01 | 0.11 |
| Tropical | 1.03 | 1.00, 1.05 | 0.054 | 1.03 | 1.00, 1.05 | **0.046** | 1.03 | 1.00, 1.05 | 0.058 | 1.01 | 0.98, 1.04 | 0.5 | 1.03 | 1.00, 1.05 | 0.059 |
| **SES Group** |  |  |  |  |  |  |  |  |  |  |  |  |  |  |  |
| Poorest | Ref | Ref |  | Ref | Ref |  | Ref | Ref |  | Ref | Ref |  | Ref | Ref |  |
| Poor | 0.93 | 0.91, 0.95 | **<0.001** | 0.93 | 0.91, 0.95 | **<0.001** | 0.93 | 0.91, 0.95 | **<0.001** | 0.93 | 0.91, 0.95 | **<0.001** | 0.93 | 0.91, 0.95 | **<0.001** |
| Middle | 0.90 | 0.87, 0.92 | **<0.001** | 0.90 | 0.87, 0.92 | **<0.001** | 0.90 | 0.87, 0.92 | **<0.001** | 0.89 | 0.87, 0.92 | **<0.001** | 0.90 | 0.87, 0.92 | **<0.001** |
| Rich | 0.86 | 0.83, 0.88 | **<0.001** | 0.86 | 0.83, 0.88 | **<0.001** | 0.86 | 0.83, 0.88 | **<0.001** | 0.86 | 0.83, 0.88 | **<0.001** | 0.86 | 0.83, 0.89 | **<0.001** |
| Richest | 0.78 | 0.74, 0.82 | **<0.001** | 0.77 | 0.73, 0.82 | **<0.001** | 0.78 | 0.74, 0.82 | **<0.001** | 0.77 | 0.73, 0.82 | **<0.001** | 0.78 | 0.74, 0.82 | **<0.001** |
| **Age of Child (Years 0-4)** | 0.78 | 0.77, 0.78 | **<0.001** | 0.78 | 0.77, 0.78 | **<0.001** | 0.78 | 0.77, 0.78 | **<0.001** | 0.78 | 0.77, 0.78 | **<0.001** | 0.78 | 0.77, 0.78 | **<0.001** |
| **STRE, 15-21 Days** | 0.96 | 0.90, 1.02 | 0.2 | 0.99 | 0.95, 1.03 | 0.6 | 1.01 | 0.97, 1.06 | 0.6 | 1.03 | 0.99, 1.07 | 0.12 | 0.98 | 0.95, 1.01 | 0.13 |
| **Animal\*STRE Interaction Term** | 1.04 | 0.97, 1.11 | 0.3 | 1.00 | 0.94, 1.05 | 0.9 | 0.96 | 0.90, 1.02 | 0.2 | 0.89 | 0.84, 0.95 | **<0.001** | 1.19 | 1.06, 1.34 | **0.003** |
| **Joint Effect from Interaction Term** | 1.00 | 0.96, 1.03 | 0.82 | 1.01 | 0.96, 1.05 | 0.78 | 0.97 | 0.94, 1.01 | 0.14 | 0.87 | 0.83, 0.92 | **<0.001** | 1.15 | 1.03, 1.29 | **0.01** |

PR = Prevalence Ratio, CI = Confidence Interval, Ref = Reference Level.
Bolded values indicate statistical significance at an $\alpha$ level of 0.05 for each individual covariate.

Additionally, in all models, the prevalence of two-week diarrhea decreased by 22% for each additional year of the child’s age (PR: 0.78, 95% CI: 0.77-0.78, p-value: <0.001). There was no statistically significant difference in the two-week prevalence of diarrhea among children that were exposed to a STRE 15-21 days before survey administration compared to children that weren’t exposed to a STRE during the same timeframe. The joint effects in our model shows no statistically significant change in the two-week diarrhea prevalence among children who se families own poultry, bull/cow/cattle, and goat/sheep, who were also exposed to an STRE in the 15-21 days before survey administration when compared to children whose families did not own these animals and were not exposed to an STRE in the same time frame. There was a statistically significant decrease in this relationship among families who owned horses or donkeys (PR: 0.87, 95% CI: 0.83-0.92, p-value: <0.001) and there was a statistically significant increase in this relationship among families who owned pigs (PR: 1.15, 95% CI: 1.03-1.29, p-value: 0.01). The results of our sensitivity analyses using the 99^th^ and 99.9^th^ percentile as the cut off for the definition of an STRE showed similar results (see supplemental tables 3 and 4).

## Discussion

The results of our analysis show that the relationship between STREs 15-21 days before survey administration and the two-week prevalence of diarrhea may be dependent on the species of animal that a household owns. Our models show that there is a decreased two-week prevalence in this relationship among families that own horses or donkeys (PR: 0.87, 95% CI: 0.83-0.92, p-value: <0.001), but an increased two-week prevalence in this relationship among families that own pigs (PR: 1.15, 95% CI: 1.03-1.29, p-value: 0.01).

In our model, the two-week prevalence of diarrhea among families who did and did not experience an STRE was mixed. Some models, such as those where poultry, bull/cow/cattle, and pigs were the primary exposures, show a point prevalence that indicates that exposure to STREs lead to a decreased prevalence of diarrhea, while models that had goat/sheep and horse/donkey as the primary exposure show a point prevalence that indicates that exposure to STREs lead to an increased prevalence of diarrhea. In all of these models, this relationship was not statistically significant.

These results are slightly different than our bivariate analyses. These analyses identified that children from families owning animals experienced a higher prevalence of diarrhea when exposed to STREs 15–21 and 22–28 days prior to the survey, compared to those not exposed to STREs during the same period. The families who did not experience STREs also had a statistically significant higher SES level compared to those who did experience STREs. This result is expected to be more of a coincidence rather than a significant finding, as there are no indications that a family’s SES level has a causal effect on the particular weather patterns that they experience.

Independent of the joint effect of STRE, children whose families own bulls, cows, or cattle have a slightly increased two-week prevalence of diarrhea compared to children whose families do not own these animals (PR: 1.02, 95% CI: 1.00-1.04, p-value: 0.04), while children whose families own horses and donkeys have a reduced prevalence of two-week diarrhea compared to households that do not own these animals (PR: 0.95, 95% CI: 0.92-0.98, p-value: <0.001), which is consistent with our previous findings.

According to our model results, the climate zone where a child lives may only have a small effect their two-week diarrhea prevalence, as the only two models with statistically significant findings were the model with bulls, cows, and cattle as the primary exposure (tropical KGZ has an increased two-week prevalence of diarrhea compared to dry KGZ) and the model with horses and donkeys as the primary exposure (temperate KGZ has a decreased two-week prevalence of diarrhea compared to dry KGZ). It’s important to note that KGZs are a proxy for seasonality in these models, and may not account for areas that have many different weather patterns in a small area, such as areas on top of mountains or down in valleys.

Previous studies have also looked at the association of EPEs on the prevalence of diarrhea in a household. A study by Bandyopadhyay et al. [17] found that heavy rainfall during dry periods in sub-Saharan Africa decreased the prevalence of diarrhea by three percent. However, this study conducted analyses differently than others, by only including diarrhea outcomes among children ages three and younger (as opposed to diarrhea outcomes among children four and younger in most other studies), and by aggregating the weather and household data to regions within each country, instead of keeping it at the cluster-level, as in most other research that uses DHS data. A study by Dimitrova et al. showed that different climates (as defined by the Köppen-Geiger) have different associations with prevalence of diarrhea depending on the current local weather patterns [18]. For example, the prevalence of diarrhea is increased during droughts in tropical savanna regions, and there is an increased prevalence of diarrhea during heavy precipitation in humid subtropical regions.

A metanalysis by Kraay et al. [6] showed that the prevalence of diarrhea increased when extreme rain was preceded by a dry period (IRR: 1.26, 95% CI: 1.05, 1.51), but not when following a moderately wet or a wet period. This metanalysis further showed that the strongest associations of diarrhea prevalence and extreme rain were between 0-2 weeks after the extreme rain event. This relationship was something that we were unable to test as part of this research, as the data from the surveys have only recently started separating whether the diarrhea occurred in the previous 24 hours or within the previous two weeks. This analysis also didn’t account for the weather before the 15-21 day lag period, and whether the STREs came after a drought or a wet period.

One strength of our study is the large number of households included in the analysis. We were able to assess the diarrheal outcomes of 413,576 children under the age of five across 99 surveys and 35 countries using nationally representative surveys. Another strength of this study is our ability to look at the different species that a household owns and their associated 2-week prevalence of diarrhea. A cross-sectional study from Uganda showed that the more animals that a family owned decreased the prevalence of diarrhea in their children under the age of 5 [19]. Previous studies have looked at the association between density of animals and nearby reported illnesses [20,21], with mixed results.

Some limitations to our study include the use of cross-sectional data, so causation cannot be ascertained using these data. With these analyses, we are only suggesting associations between aspects of the data. Additionally, a household could only state up to a maximum of 95 of a type of animal. This most severely limits our information regarding the true ownership patterns of chickens, ducks, and other poultry among households, as these are the cheapest animals to own compared to the other four categories of animals we examined in terms of both cost and care [3]. Another limitation was the lack of information about where these animals were housed. Chickens may be raised closer to the home compared to other larger livestock, which may expose young children to more chicken fecal pathogens compared to other animals’ fecal pathogens [22,23]. More information about animal husbandry practices is needed to more fully understand the complete relationship between animal ownership and prevalence of diarrhea, especially during periods of extreme precipitation.

Another limitation for this study is the reporting of diarrhea by the primary caregiver. It has been well-documented that the reporting of diarrhea by the primary caregiver can be flawed [24–26], particularly in DHS surveys [27]. Additionally, exposure to animal pathogens is just a single pathway that may lead to an incidence of diarrhea. A metanalysis found that among eight studies, exposure to fecal pathogens of animal origin may or may not impair child growth [28], bringing into question whether a focus on fecal pathogens in the environment is a necessary focus.

Future studies can further refine the use of STREs in their analyses. Our analysis used a binary definition of whether an STRE occurred within a specific timeframe. However, it would be valuable to know of the local weather patterns before the 15-21 day lag period used in this analysis, and whether that weather had already diluted the environment of diarrheal pathogens. Our definition of an STRE also depends on the time of the year. With our definition, which is based on a precipitation event that is above the 95^th^ percentile, a slight precipitation during a very dry period would count as an STRE, but may not be enough to transport pathogens in the environment. On the opposite side of the spectrum, during a wet period, the threshold for an STRE is higher. Even though there’s a lot of precipitation in the environment that could transport pathogens, an increase in precipitation would not necessarily register as an STRE. Time frames are also important to consider when conducting surveys, as animal husbandry practices differ depending on religious (e.g. Eid al’Adha) and non-religious (e.g. Independence Day) celebrations. People may also shift their husbandry practices by moving animals if they expect a lot of rain or other adverse weather in the area. Smaller studies focused on smaller cohorts may be able to take advantage of these considerations in the design of their surveys.

This research has sought to expand the discourse about the concentration-dilution hypothesis by expanding the focus to peridomestic animals throughout sub-Saharan Africa. We have shown that ownership of some animals, such as horses, donkeys, and pigs, may have an effect of the prevalence of diarrhea when also considering local STREs. Improved management of animal feces, especially among animals that live close to houses and compounds, may have an effect on the diarrhea prevalence of children under the age of five. This should be an increased focus when STREs are expected after increased dry periods.

## Data Availability

All data produced in the present study are available upon reasonable request to the authors

## Acknowledgements

This work was carried out at the Advanced Research Computing at Hopkins (ARCH) core facility (rockfish.jhu.edu), which is supported by the National Science Foundation (NSF) grant number OAC1920103.

---

**Supplement Table 1.**
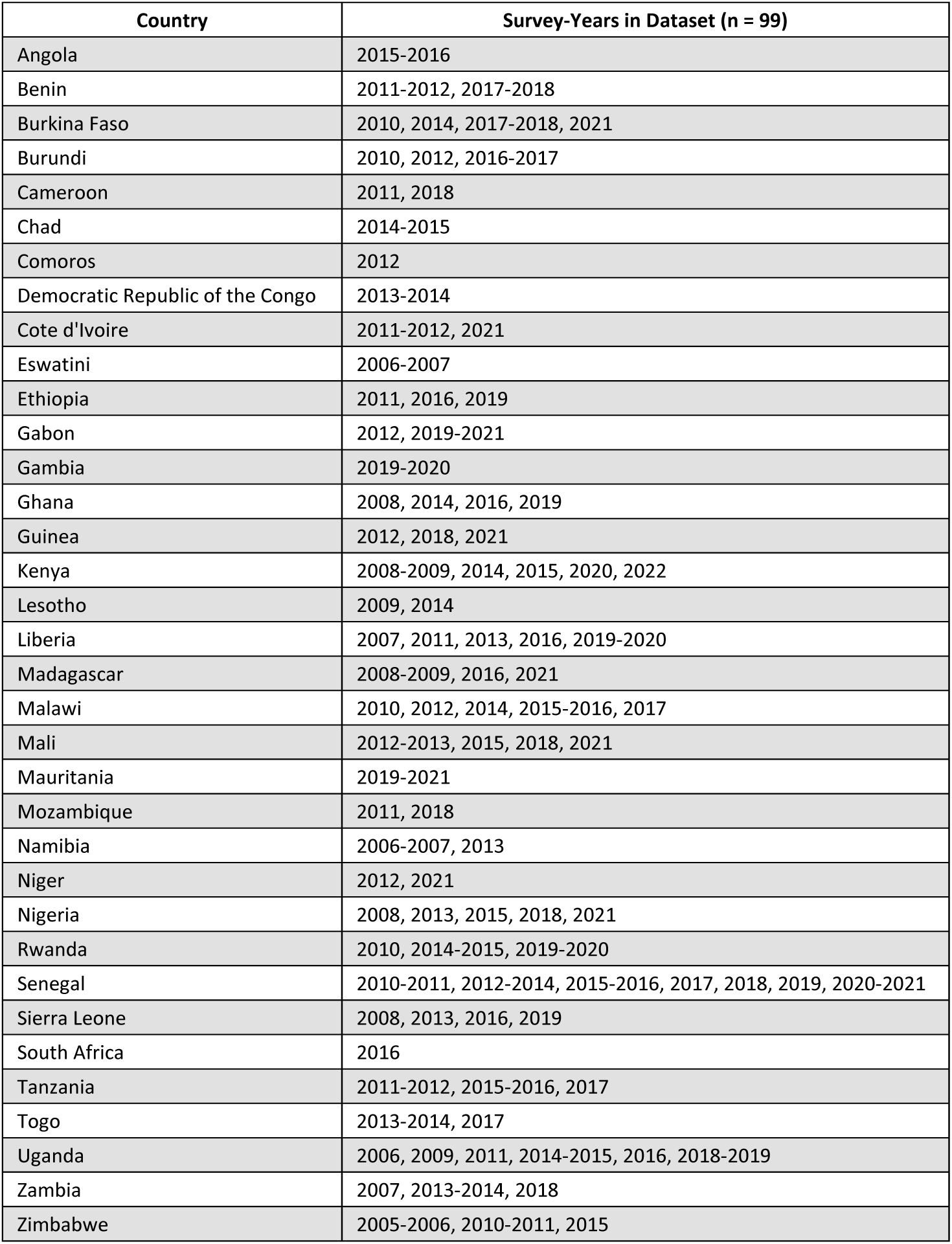
Countries and survey-years used in this analysis

| Country | Survey-Years in Dataset (n = 99) |
| --- | --- |
| Angola | 2015-2016 |
| Benin | 2011-2012, 2017-2018 |
| Burkina Faso | 2010, 2014, 2017-2018, 2021 |
| Burundi | 2010, 2012, 2016-2017 |
| Cameroon | 2011, 2018 |
| Chad | 2014-2015 |
| Comoros | 2012 |
| Democratic Republic of the Congo | 2013-2014 |
| Cote d'Ivoire | 2011-2012, 2021 |
| Eswatini | 2006-2007 |
| Ethiopia | 2011, 2016, 2019 |
| Gabon | 2012, 2019-2021 |
| Gambia | 2019-2020 |
| Ghana | 2008, 2014, 2016, 2019 |
| Guinea | 2012, 2018, 2021 |
| Kenya | 2008-2009, 2014, 2015, 2020, 2022 |
| Lesotho | 2009, 2014 |
| Liberia | 2007, 2011, 2013, 2016, 2019-2020 |
| Madagascar | 2008-2009, 2016, 2021 |
| Malawi | 2010, 2012, 2014, 2015-2016, 2017 |
| Mali | 2012-2013, 2015, 2018, 2021 |
| Mauritania | 2019-2021 |
| Mozambique | 2011, 2018 |
| Namibia | 2006-2007, 2013 |
| Niger | 2012, 2021 |
| Nigeria | 2008, 2013, 2015, 2018, 2021 |
| Rwanda | 2010, 2014-2015, 2019-2020 |
| Senegal | 2010-2011, 2012-2014, 2015-2016, 2017, 2018, 2019, 2020-2021 |
| Sierra Leone | 2008, 2013, 2016, 2019 |
| South Africa | 2016 |
| Tanzania | 2011-2012, 2015-2016, 2017 |
| Togo | 2013-2014, 2017 |
| Uganda | 2006, 2009, 2011, 2014-2015, 2016, 2018-2019 |
| Zambia | 2007, 2013-2014, 2018 |
| Zimbabwe | 2005-2006, 2010-2011, 2015 |

**Supplemental Table 2.** Common Pathogens in Africa and their associated incubation periods. Pathogen Illnesses and Deaths are taken from a study by Thystrup et al. (2024).

| Pathogen | Type | Estimated Incubation Time | Whole African Region (197 mil. Pop) |  |
| --- | --- | --- | --- | --- |
|  |  |  | Illnesses (95% UI) | Deaths (95% UI) |
| Rotavirus | Virus | 1-2 Days ^ | 9,457,547 (4,931,180–13,115,905) | 27,616 (18,226–45,459) |
| Norovirus | Virus | 12-48 Hours ^ | 10,257,653 (4,650,470–14,555,680) | 9,135 (5,980–15,005) |
| Astrovirus | Virus | 4-5 days ^ | 2,544,662 (1,196,629-3,590,575) | 457 (299–750) |
| Cryptosporidium spp. | Protozoan | 2-10 days (usually 7 days) § | 5,901,845 (2,695,492-8,365,420) | 22,121 (14,525–36,363) |
| Entamoeba histolytica | Protozoan | 2-4 weeks § | 883,095 (418,536-1,244,527) | 1,802 (1,161-2,948) |
| Giardia lamblia | Protozoan | 1-14 days (usually 7 days) § | 14,037,258 (6,437,064–19,884,469) | 9,198 (6,079–15,148) |
| Campylobacter spp. | Bacteria | 1-10 days (usually 2-4 days) § | 5,902,020 (2,611,903-8,405,257) | 8,108 (5,271–13,293) |
| Enterohemorrhagic E. coli (EHEC): | Bacteria | 2-5 days & | 2,919,966 (1,266,400-4,170,655) | 5,076 (3,290-8,315) |
| Enteropathogenic E. coli (EPEC): | Bacteria | 9-12 hours § | 7,517,917 (3,370,830–10,685,772) | 12,317 (8,037–20,214) |
| Enterotoxigenic E. coli (ETEC): | Bacteria | 10-72 hours§ | 10,044,953 (4,476,717–14,290,494) | 6,988 (4,607–11,500) |
| Enteraggregative E. coli (EAEC): | Bacteria | 8-48 hours§ | 17,137,586 (7,924,333–24,245,224) | 21,471 (14,013–35,237) |
| Enteroinvasive E. coli (EIEC): | Bacteria | 10-18 hours§ | 3,961,764 (2,027,562-5,512,315) | 70,108 (46,172–115,341) |
| Shiga toxin-producing E. coli (STEC): | Bacteria | 1–10 days (usu. 3–4 days) § | 8,468,956 (4,086,157–11,900,853) | 633 (428-1,050) |
| V. cholerae | Bacteria | 1-2 days* | 76,033 (43,923–103,420) | 3,968 (2,697-6,586) |
| Salmonella spp. | Bacteria | 12 hours - 4 days^ | 889,172 (422,620-1,252,524) | 700 (479-1,163) |
| Shigella spp. | Bacteria | 1-2 days§ | 2,050,246 (971,448-2,889,486) | 10,596 (6,929–17,398) |

^ Source: Cleveland Clinic
§ Source: CDC Yellow Book
\* Azman et al., 2014
& Hopkins Medicine

**Supplemental Table 3.** Model results from the mixed-effects Poisson regression models with a log link. The outcome of these models is a prevalence ratio of children with two-week diarrhea compared to children that did not experience two-week diarrhea. An STRE as a single day of precipitation that was above the 99^th^ percentile of the total precipitation of the time period.

|  | Poultry (Chickens/Ducks) |  |  | Bull/Cow/Cattle |  |  | Goat/Sheep |  |  | Horse/Donkey |  |  | Pig |  |  |
| --- | --- | --- | --- | --- | --- | --- | --- | --- | --- | --- | --- | --- | --- | --- | --- |
|  | PR | 95% CI | p-value | PR | 95% CI | p-value | PR | 95% CI | p-value | PR | 95% CI | p-value | PR | 95% CI | p-value |
|  | 1.00 | 0.98, 1.02 | 0.9 |  |  |  |  |  |  |  |  |  |  |  |  |
|  |  |  |  | 1.02 | 1.00, 1.05 | **0.03** |  |  |  |  |  |  |  |  |  |
|  |  |  |  |  |  |  | 1.00 | 0.98, 1.02 | 0.9 |  |  |  |  |  |  |
|  |  |  |  |  |  |  |  |  |  | 0.95 | 0.93, 0.98 | **0.003** |  |  |  |
|  |  |  |  |  |  |  |  |  |  |  |  |  | 0.99 | 0.96, 1.02 | 0.5 |
| Köppen-Geiger Zone |  |  |  |  |  |  |  |  |  |  |  |  |  |  |  |
| Dry | Ref | Ref |  | Ref | Ref |  | Ref | Ref |  | Ref | Ref |  | Ref | Ref |  |
| Temperate | 0.97 | 0.93, 1.01 | 0.12 | 0.97 | 0.93, 1.01 | 0.10 | 0.97 | 0.93, 1.01 | 0.12 | 0.96 | 0.92, 1.00 | **0.028** | 0.97 | 0.93, 1.01 | 0.12 |
| Tropical | 1.03 | 1.00, 1.05 | 0.057 | 1.03 | 1.00, 1.06 | **0.045** | 1.03 | 1.00, 1.05 | 0.051 | 1.01 | 0.98, 1.04 | 0.5 | 1.03 | 1.00, 1.05 | 0.056 |
| SES Group |  |  |  |  |  |  |  |  |  |  |  |  |  |  |  |
| Poorest | Ref | Ref |  | Ref | Ref |  | Ref | Ref |  | Ref | Ref |  | Ref | Ref |  |
| Poor | 0.93 | 0.91, 0.95 | **<0.001** | 0.93 | 0.91, 0.95 | **<0.001** | 0.93 | 0.91, 0.95 | **<0.001** | 0.93 | 0.91, 0.95 | **<0.001** | 0.93 | 0.91, 0.95 | **<0.001** |
| Middle | 0.90 | 0.87, 0.92 | **<0.001** | 0.90 | 0.87, 0.92 | **<0.001** | 0.90 | 0.87, 0.92 | **<0.001** | 0.89 | 0.87, 0.92 | **<0.001** | 0.90 | 0.87, 0.92 | **<0.001** |
| Rich | 0.86 | 0.83, 0.88 | **<0.001** | 0.86 | 0.83, 0.88 | **<0.001** | 0.86 | 0.83, 0.88 | **<0.001** | 0.86 | 0.83, 0.88 | **<0.001** | 0.86 | 0.83, 0.89 | **<0.001** |
| Richest | 0.78 | 0.74, 0.82 | **<0.001** | 0.78 | 0.74, 0.82 | **<0.001** | 0.78 | 0.74, 0.82 | **<0.001** | 0.77 | 0.73, 0.82 | **<0.001** | 0.78 | 0.74, 0.82 | **<0.001** |
| Age of Child (Years 0-4) | 0.78 | 0.77, 0.78 | **<0.001** | 0.78 | 0.77, 0.78 | **<0.001** | 0.78 | 0.77, 0.78 | **<0.001** | 0.78 | 0.77, 0.78 | **<0.001** | 0.78 | 0.77, 0.78 | **<0.001** |
| STRE, 15-21 Days | 0.95 | 0.89, 1.01 | 0.12 | 1.00 | 0.96, 1.05 | >0.9 | 1.02 | 0.97, 1.08 | 0.4 | 1.05 | 1.01, 1.10 | **0.015** | 0.98 | 0.95, 1.01 | 0.3 |
| Animal*EPE Interaction Term | 1.06 | 0.98, 1.13 | 0.13 | 0.98 | 0.92, 1.04 | 0.5 | 0.96 | 0.90, 1.02 | 0.2 | 0.86 | 0.81, 0.92 | **<0.001** | 1.22 | 1.08, 1.39 | **0.002** |
| Joint Effect from Interaction Term | 1.00 | 0.96, 1.05 | 0.85 | 1.00 | 0.96, 1.05 | 0.86 | 0.98 | 0.94, 1.02 | 0.24 | 0.86 | 0.82, 0.91 | **<0.001** | 1.19 | 1.05, 1.34 | **0.006** |

PR = Prevalence Ratio, CI = Confidence Interval, Ref = Reference Level.
Bolded values indicate statistical significance at an $\alpha$ level of 0.05 for each individual covariate.

**Supplemental Table 4.** Model results from the mixed-effects Poisson regression models with a log link. The outcome of these models is a prevalence ratio of children with two-week diarrhea compared to children that did not experience two-week diarrhea. An STRE as a single day of precipitation that was above the 99.9^th^ percentile of the total precipitation of the time period.

|  | Poultry (Chickens/Ducks) |  |  | Bull/Cow/Cattle |  |  | Goat/Sheep |  |  | Horse/Donkey |  |  | Pig |  |  |
| --- | --- | --- | --- | --- | --- | --- | --- | --- | --- | --- | --- | --- | --- | --- | --- |
|  | PR | 95% CI | p-value | PR | 95% CI | p-value | PR | 95% CI | p-value | PR | 95% CI | p-value | PR | 95% CI | p-value |
|  | 1.00 | 0.97, 1.02 | 0.8 |  |  |  |  |  |  |  |  |  |  |  |  |
|  |  |  |  | 1.02 | 1.00, 1.04 | **0.03** |  |  |  |  |  |  |  |  |  |
|  |  |  |  |  |  |  | 1.00 | 0.98, 1.02 | 0.8 |  |  |  |  |  |  |
|  |  |  |  |  |  |  |  |  |  | 0.95 | 0.93, 0.98 | **0.002** |  |  |  |
|  |  |  |  |  |  |  |  |  |  |  |  |  | 0.99 | 0.96, 1.02 | 0.6 |
| **Köppen-Geiger Zone** |  |  |  |  |  |  |  |  |  |  |  |  |  |  |  |
| Dry | Ref | Ref |  | Ref | Ref |  | Ref | Ref |  | Ref | Ref |  | Ref | Ref |  |
| Temperate | 0.97 | 0.93, 1.01 | 0.12 | 0.97 | 0.93, 1.01 | 0.10 | 0.97 | 0.93, 1.01 | 0.12 | 0.96 | 0.92, 1.00 | **0.029** | 0.97 | 0.94, 1.01 | 0.12 |
| Tropical | 1.03 | 1.00, 1.05 | 0.055 | 1.03 | 1.00, 1.06 | **0.043** | 1.03 | 1.00, 1.05 | 0.052 | 1.01 | 0.98, 1.04 | 0.5 | 1.03 | 1.00, 1.05 | 0.054 |
| **SES Group** |  |  |  |  |  |  |  |  |  |  |  |  |  |  |  |
| Poorest | Ref | Ref |  | Ref | Ref |  | Ref | Ref |  | Ref | Ref |  | Ref | Ref |  |
| Poor | 0.93 | 0.91, 0.95 | **<0.001** | 0.93 | 0.91, 0.95 | **<0.001** | 0.93 | 0.91, 0.95 | **<0.001** | 0.93 | 0.91, 0.95 | **<0.001** | 0.93 | 0.91, 0.95 | **<0.001** |
| Middle | 0.90 | 0.87, 0.92 | **<0.001** | 0.90 | 0.87, 0.92 | **<0.001** | 0.90 | 0.87, 0.92 | **<0.001** | 0.89 | 0.87, 0.92 | **<0.001** | 0.90 | 0.87, 0.92 | **<0.001** |
| Rich | 0.86 | 0.83, 0.88 | **<0.001** | 0.86 | 0.83, 0.88 | **<0.001** | 0.86 | 0.83, 0.88 | **<0.001** | 0.86 | 0.83, 0.88 | **<0.001** | 0.86 | 0.83, 0.89 | **<0.001** |
| Richest | 0.78 | 0.74, 0.82 | **<0.001** | 0.78 | 0.74, 0.82 | **<0.001** | 0.78 | 0.74, 0.82 | **<0.001** | 0.77 | 0.73, 0.82 | **<0.001** | 0.78 | 0.74, 0.82 | **<0.001** |
| **Age of Child (Years 0-4)** | 0.78 | 0.77, 0.78 | **<0.001** | 0.78 | 0.77, 0.78 | **<0.001** | 0.78 | 0.77, 0.78 | **<0.001** | 0.78 | 0.77, 0.78 | **<0.001** | 0.78 | 0.77, 0.78 | **<0.001** |
| **STRE, 15-21 Days** | 0.95 | 0.89, 1.01 | 0.11 | 1.01 | 0.96, 1.05 | 0.8 | 1.02 | 0.97, 1.08 | 0.5 | 1.06 | 1.02, 1.10 | **0.008** | 0.99 | 0.95, 1.02 | 0.4 |
| **Animal\*EPE Interaction Term** | 1.07 | 0.99, 1.15 | 0.08 | 0.98 | 0.92, 1.04 | 0.5 | 0.96 | 0.90, 1.03 | 0.3 | 0.86 | 0.80, 0.92 | **<0.001** | 1.22 | 1.07, 1.40 | **0.004** |
| **Joint Effect from Interaction Term** | 1.01 | 0.97, 1.05 | 0.63 | 1.01 | 0.96, 1.06 | 0.71 | 0.98 | 0.94, 1.02 | 0.39 | 0.87 | 0.82, 0.92 | **<0.001** | 1.19 | 1.05, 1.35 | **0.01** |

PR = Prevalence Ratio, CI = Confidence Interval, Ref = Reference Level.
Bolded values indicate statistical significance at an $\alpha$ level of 0.05 for each individual covariate.

